# Relative cost of counselling compared to supplementary foods for acute malnutrition: a time and motion study in Nairobi Kenya

**DOI:** 10.1101/2023.08.27.23294692

**Authors:** Antonina N. Mutoro, Eleanor Grieve, Ada L. Garcia, Charlotte Wright

**Affiliations:** Human Nutrition department, School of Medicine, Dentistry and Nursing, University of Glasgow UK; Health Economics and Health Technology Assessment, School of Health and Wellbeing, University of Glasgow, UK; Maternal and Child Wellbeing Unit, African Population and Health Research Centre (APHRC) Nairobi, Kenya

## Abstract

**Aims:** We aimed to describe the relative cost of supplying supplementary food, compared to counselling, in children with acute malnutrition managed in one urban area.

**Method:** A cross-sectional costing study was undertaken. Interviews with staff and time-and-motion observations of children with acute malnutrition and their carers attending 6 ambulatory nutrition treatment centres in Nairobi were conducted to assess the time nutritionists spent treating acute malnutrition and managing ready to use food supplies. Kenyan standard pay rates ($) were used to calculate the monthly staff cost of reviewing each child and speaking to their parent. These were compared to the published cost of purchasing daily ready to use food sachets.

**Results:** Consultations with 32 children were observed, 3-8 per clinic. Staff spent mean (SD) 9.84 (4.9) minutes seeing each child, of which at most only 3.5 (2.0) minutes were spent counselling the family. With an hourly pay rate of $4.87, the median (IQR) contact and dispensing cost per child visit was $0.77 (0.49-1.16). The cost of RUF was estimated to be $7.84 for a month’s treatment, giving a total treatment cost of $8.61 per child per month. If ready to use food was not prescribed and more staff time purchased instead, this would pay for 1.6 hours staff time per child per month.

**Conclusions:** Very little time was spent speaking to mothers of malnourished children and supplies of ready to use food represented 91% of total treatment costs.

## Introduction

Moderate acute malnutrition (MAM) is a major public health problem in many low- and middle-income countries (LMIC). It is accountable for more deaths in children under 5 years than severe acute malnutrition (SAM) [1]. In 2019, approximately 32.7 million children were moderately wasted [2]. However, the best approach to treatment of MAM and rehabilitation of SAM after initial rescue remains unclear. Many supplementary feeding interventions have been trialled which aim to correct energy and nutrient deficiencies using specially formulated foods such as blended flours or, more recently, ready-to-use foods (RUF) which are offered in addition to a child’s normal diet [3], but simply giving supplementary food has only limited effects [4[5] and their use is still not recommended by WHO, except in emergency settings. [6] This limited efficacy may reflect the multiple possible interacting causes of child undernutrition [7] and the possibility of misdiagnosis of intrauterine stunting as current undernutrition [6]. Nonetheless RUF is already used in the treatment of MAM in many countries, including Kenya [8]. At present RUF supplies are generally donated by external agencies, which presents sustainability challenges and recurrent periods with no supplies. As well as supplying RUF, the Kenyan MAM protocol specifies a baseline assessment, which should identify proximal issues, such as diet and caring practises, with nutritional and welfare advice given as required. However, this requires time, and most treatment centres are already heavily burdened and understaffed. We are developing an algorithm for the assessment of children with MAM to ensure that only acutely wasted children are identified as MAM using a proforma that ensures that all relevant risk factors are recorded and can inform advice and support for the family [9]. However, this takes around 20 minutes to complete, with further time needed at follow-up to reinforce advice given. In order to plan a trial of its use, we needed to know how much time is currently spent providing RUF and how much it would cost to fund extra staff time for counselling. This assessment would be expected to result in less RUF being prescribed overall, so we also considered how much staff time could be funded by a reduction in expenditure on RUF. Previous economic studies have tended only to examine the cost effectiveness of community management of SAM [10-12] or to compare the cost of different RUF used in MAM without untreated or counselling comparisons[13]. We thus planned an exploratory time and motion study to establish cost of staff time spent with families and administering and dispensing RUF, as well as the cost of RUF itself.

## Methods

### Study design and setting

This cross-sectional costing study was conducted in 2019 in Nairobi in 5 government health centres and one faith-based health facility of the 80 facilities in Nairobi which offer ambulatory therapeutic (OTP) and supplementary feeding programs (SFP) for acute malnutrition, (severe or moderate). These centres were pragmatically selected because they served informal settlements which have high malnutrition rates [14, 15]. Interviews and observations (time and motion studies) were used to ascertain time spent delivering and administering treatment.

### Ethics approval

Permission to conduct the study was obtained from the African Medical and Research Foundation ethics review committee, and the county and sub county health offices. Study participants were recruited between January and March 2019. Written informed consent was obtained from participants before recruitment. The research team approached mothers, provided information about the study and inquired if they were willing to participate in the study. Participants who were willing to participate were requested to sign two copies of a consent form one of which they kept. The other copy was left with the researchers.

### Participants

The subjects of the time and motion observations were children and their carers attending OTP and SFP clinics and the health workers who were actively involved in their treatment. The sessions were held within child welfare clinics and reviewed both children with MAM and uncomplicated SAM, so both were included. MAM was defined as weight for length Z scores (WLZ) between -2SD and -3SD and SAM WLZ≤-3SD. Children who had complications that required specialised care were referred elsewhere and excluded from the study.

### Interviews

For the interview element the health worker responsible for MAM/SAM in each health facility was identified and interviewed using a structured schedule to establish direct and indirect costs incurred during the treatment process. Each participant was interviewed once. Information was collected about the grade of staff seeing malnourished children and matched to published pay scales [16]. They estimated the total number of children seen per day as well as time spent by staff on requisition and management of RUF and in relevant training; and RUF transportation costs were also collected.

### Time and motion observations

The same researchers attended each of the 6 facilities to observe one clinic sessions in each. During each observed clinic mothers of children attending for MAM/SAM treatment were identified on arrival and observed by the researcher throughout their attendance on that day. For each child key activities were recorded, and the time taken for each noted:

1. Anthropometric assessments (Anthropometry)
2. Collection and recording of patient information (History taking)
3. Appetite test
4. Treatment prescription, counselling/advising (Counselling)
5. RUF management (RUF)

If a child required other services such as treatment for other ailments, the research assistant would pause the clock and resume timing once the child was back for OTP/SFP services. The total length of time spent on each child was then recorded.

### Cost of RUF

The RUF were donated rather than purchased, so that the actual cost of supplies was unknown. We thus estimated the cost based on information published for by UNICEF for 2019 [17]

### Analysis

Stata Version 17 was used for the analysis. The two outcomes of interest were the total monthly staff cost of reviewing each child and dispensing RUF compared to the cost of purchasing daily RUF sachets. An ingredients-based approach was used to collect staff costs, whereby each resource required was identified and valued and healthcare provider’s costs were considered. The staff costs of two main activities were considered: treatment of children with acute malnutrition and administration of RUF. The healthcare premises were treated as a standard capital cost, as any variation of interest was in the staffing time and resource use. The total time spent by each staff member per quarter for the planning and requisition for the RUF reported at the interview was recorded and converted to monthly (and annual) costs.

The cost related to the planning and requisition for the RUF was the total time spent per month in hours by nutritionists in these activities, multiplied by the hourly wage. Data on time spent in training related to the planning and requisition of RUF and transport costs were also added.

From the observational survey, we knew the average time spent per child per visit and the average per MAM and SAM child. From the interviews we knew the total number of children under treatment and the total amount of time per quarter spent on administering and dispensing RUF, so we could calculate the average cost per child on these central activities. The cost of nutritionists was calculated using their hourly wage. All staff costs were reported in US dollars and 2019 is used as the base year for the costing. At that time KES 1= USD 0.0098.

## Results

### Time and motion results

Thirty-two children were observed, with 3-8 children observed per clinic; 20 were being treated for MAM and 12 for SAM. Ages ranged from 7 to 26 months, with a mean (SD) of 13 (4.3) months. For 6 children, this was their first appointment with the rest attending follow-up clinics.

In 3 of the centres the pre-stated activities were often observed as happening at the same time, so that the amount of time spent on each activity could not be determined, except for anthropometry, but counselling was recorded as happening as a sole or mixed activity in 29 (90%) children; history taking was observed in only 4 (12%) children.

The median (range) time spent by the family at the facility was 15 (5-46) minutes, but the mean (SD) time in all nutrition related activities was only 9.84 (5.37) minutes, of which 2.7 (1.6) minutes was spent being measured (Table1). If one counts all the time where activity 4 was recorded within a sole or mixed activity as counselling, the maximum time spent counselling was 3.5 (2.0) minutes and only slightly more time was spent with children with SAM.

**Table 1:** Time and motion results: time spent in different activities broken down by severity.

| Time spent (minutes) on | Anthropometry | Counselling, including when mixed with other activities | Total contact time |
| --- | --- | --- | --- |
| Undernutrition status | Mean (SD) | Mean (SD) | Mean (SD) |
| Moderate acute malnourished cases (n=20) | 2.3 (1.2) | 3.9 (1.5) | 8.6 (4.8) |
| Severe acute malnourished cases (n=12) | 3.3 (2.2) | 3.1 (2.5) | 11.9 (4.7) |
| Total (n=32) | 2.69 (1.6) | 3.55 (2.0) | 9.84 (4.9) |

### Staff costs

The costs related to management of undernutrition and estimate the monthly average cost per child contact are shown in Table 2. Centres saw on average 28 children a day (SAM and MAM combined), but this varied widely (range 7-80 children per clinic). The hourly pay for nutritionists was $4.87 [16] resulting in a median cost per child contact visit of $0.73, a dispensing cost $0.03 and total cost of $0.77.

**Table 2:** Monthly costs per child, per contact.

| Centre | Number contacts per month | Total costs children contact (\$) | Dispensing Costs (\$) | Costs per child (\$) | | |
| --- | --- | --- | --- | --- | --- | --- |
|  |  |  |  | Contact | Dispensing | Total |
| 1 | 60 | 43.20 | 1.33 | 0.72 | 0.02 | 0.74 |
| 2 | 80 | 92.80 | 1.67 | 1.16 | 0.02 | 1.18 |
| 3 | 400 | 344.00 | 10.03 | 0.86 | 0.03 | 0.89 |
| 4 | 120 | 99.60 | 2.15 | 0.83 | 0.02 | 0.85 |
| 5 | 140 | 182.00 | 47.93 | 1.3 | 0.34 | 1.64 |
| 6 | 1171 | 597.21 | 41.45 | 0.51 | 0.04 | 0.55 |
|  |  |  | Mean | 0.82 | 0.06 | 0.87 |
|  |  |  | (95% CI) | (0.66, 0.97) | (0.02, 0.09) | (0.70, 1.05) |
|  |  |  | Median | 0.73 | 0.03 | 0.77 |
|  |  |  | (IQR) | (0.45, 1.10) | (0.02, 0.04) | (0.49, 1.16) |

Starting (first) appointments lasted significantly longer (17.5 mins) than follow-up appointments (8.35 mins), a mean difference of 9.15 95% CI (5.4, 12.9) min P <0.001.

### Cost of RUF

The UNICEF website listed one Kenyan supplier in 2019 with a cost per carton for 150 sachets of $42, which translates into a monthly cost for treatment of MAM (30 sachets) of £7.84 per child. If RUF was not prescribed and more staff time purchased instead, this would pay for 1.6 staff hours per child per month.

## Discussion

While Kenyan MAM protocols specify a targeted counselling approach in addition to provision of RUF, we found that in fact very little time was spent interacting with the parents of children with either MAM or SAM. The monthly staff cost of $0.77 per month contrasts strikingly with the $7.84 cost of the therapeutic foods dispensed which represented 91% of the total monthly treatment costs ($8.61) per child for acute malnutrition in this setting. A limitation is that the exact product costs remain uncertain, however, the best estimate of the local price appears representative of RUF prices worldwide. The low staff costs found partly reflect the small amount of time spent, but also that salary rates in Kenya are relatively low compared to more developed countries; for example compared to a median monthly income in the USA of $6,966, the median monthly salary in Kenya is $897 [18]. We found that the opportunity cost of RUF per month was equivalent to 1.6 hours of staff time, so that a reduction in RUF use by half could in principle allow staff to spend 48 more minutes per month working with families: five times the amount of time spent currently. It must also be borne in mind that salary rates in Kenya are relatively high compared to other middle income countries; for example the median monthly income in Nigeria is only $446 [18], making the opportunity cost higher in other countries.

The economics of RUF supply are complex, as the product is usually paid for by donor agencies, while staff time spent with families comes out of local health and nutrition budgets. However, at a future time when the Kenyan health systems is expected to fund RUF themselves, planner would have to consider whether RUF was a ‘better buy’ than 1.6 hours of staff time. It must also be borne in mind that fluctuations in exchange rates would mean that the cost of RUF, purchased in dollars, could be higher. At the time of submission, the dollar exchange rate is only two thirds what it was in 2019. While RUF has been shown to be more effective than standard care in the treatment of MAM, these effects are very modest; one systematic review found a net gain of only 120g in weight following treatment [4] while another found only a 10% increase in those recovering and 0.2 SD increase in weight-for-height [5]. This small overall effect may reflect only some children responding well to RUF. Screening for MAM using MUAC also identifies stunted, non-wasted children [19] who are not malnourished and could even be harmed by RUF [6]. Further research is needed to establish which children benefit most from RUF and which children can be safely left on family diet only.

Counselling has generally been found to be less effective than RUF, [20] but in most trials the counselling provided was brief or group-based and not individualised. A recent study in Kenya found many potentially reversible risk factors present in children with MAM [7] which will remain unrecognised in a programme where only RUF was offered. One recent large trial found that well-staffed individualised counselling was as effective as RUF, after allowance for higher rates of attrition [21]. Future trials of counselling will also need to consider how to incentivise parental engagement when supplementary food is not given.

Previous studies of the cost effectiveness of community management of SAM argued that they were cost effective because of the relatively low base case cost (US$53) per disability-adjusted life year gained [11] although another accepted that the worst case cost might be as much as ten times that [12]. However, the same sort of scrutiny has not yet been applied to MAM. One study compared the cost of different supplementary foods used for MAM treatment and found that their costs per recovered child were similar (US$163-79).

However, with no control group, many of these recoveries would have occurred spontaneously [22] and with no counselling arm it was not clear how this would compare to counselling [13]. The review above [5] would suggest that 10 children would neeed to be treated for one extra recovery, suggesting a cost per extra recovery possibly ten times higher. Using Kenyan salary scales this could pay a nutritionist for 2 months.

The strength of this study is that it provides a first proper disaggregation of time and product in the management of acute malnutrition. A limitation is that it examines this in only one setting. In some cases, there was an overlap of activities, so that counselling and RUF management time might also have included recording patient information, but the total time spent is still very brief. One of the included centres is known to schedule in a single counselling session separately at the end of treatment, which was not counted in this observation, and this centre had the lowest contact time overall. While the focus of this paper is MAM, we also observed the SAM children seen in the same sessions and included them in our calculations, as in practice there were only margin differences in the time spent with them.

It is often assumed that programs that require extra staff time are less affordable where a product can simply be provided. However, in an LMIC setting, staff salaries are low and even apparently cheap products prove expensive if given over longer periods. In the long term, investment in staff and skills is more likely to be sustainable and effective, as staff can address multiple health and wellbeing issues at the same time.

## Conclusions

This study reveals that very little time is currently spent speaking to mothers of malnourished children and that RUF is extremely expensive relative to other health system costs. Well-staffed trials of the efficacy and cost-effectiveness of problem-oriented counselling interventions are needed, but this study illustrates that these could be highly affordable.

## Data Availability

All relevant data are within the manuscript and its Supporting Information files.

## Acknowledgments

We are grateful to the staff of the participating clinics and to Hermann Donfouet for his initial analysis of the data.

## References

1. Olofin I, McDonald CM, Ezzati M, Flaxman S, Black RE, Fawzi WW, et al. Associations of Suboptimal Growth with All-Cause and Cause-Specific Mortality in Children under Five Years: A Pooled Analysis of Ten Prospective Studies. PLOS ONE. 2013;8(5):e64636. doi: 10.1371/journal.pone.0064636.

2. UNICEF, WHO, The World Bank. Levels and trends in child malnutrition: Key Findings of the 2020 Edition of the Joint Child Malnutrition Estimates. Geneva: World Health Organization 2020.

3. WHO. Technical Note: Supplementary foods for the management of moderate acute malnutrition in infants and children 6-59 months of age. Geneva: WHO, 2012.

4. Kristjansson E, Francis DK, Liberato S, Benkhalti Jandu M, Welch V, Batal M, et al. Food supplementation for improving the physical and psychosocial health of socio-economically disadvantaged children aged three months to five years. The Cochrane database of systematic reviews. 2015;(3):CD009924. Epub 2015/03/06. doi: 10.1002/14651858.CD009924.pub2. PubMed PMID: 25739460; PubMed Central PMCID: PMCPMC6885042.

5. Lazzerini M, Rubert L, Pani P. Specially formulated foods for treating children with moderate acute malnutrition in low- and middle-income countries. Cochrane Database of Systematic Reviews. 2013;(6). doi: 10.1002/14651858.CD009584.pub2. PubMed PMID: CD009584.

6. WHO. Guideline: assessing and managing children at primary health-care facilities to prevent overweight and obesity in the context of the double burden of malnutrition.. Geneva: World Health organization 2017.

7. Mutoro AN, Garcia AL, Kimani-Murage EW, Wright CM. Prevalence and overlap of known undernutrition risk factors in children in Nairobi Kenya. Maternal & child nutrition. 2021:e13261. Epub 2021/08/07. doi: 10.1111/mcn.13261. PubMed PMID: 34355500.

8. Kenya Ministry of Medical Services and Ministry of Public Health and Sanitation. Handbook -integrated management of acute malnutrition. 2010.

9. Mayén VA, Kimani-Murage E, Bryant-Waugh R, Traynor O, Milligan B, Khan A, et al. Are Malnourished Children Hungry? Use of the International Complementary Feeding Assessment Tool (ICFET) to Describe Diet and Eating Behavior. Current Developments in Nutrition. 2020;4(Supplement_2):924–. doi: 10.1093/cdn/nzaa053_129.

10. Garg CC, Mazumder S, Taneja S, Shekhar M, Mohan SB, Bose A, et al. Costing of three feeding regimens for home-based management of children with uncomplicated severe acute malnutrition from a randomised trial in India. BMJ Global Health. 2018;3(2):e000702. doi: 10.1136/bmjgh-2017-000702.

11. Bachmann MO. Cost-effectiveness of community-based treatment of severe acute malnutrition in children. Expert Rev Pharmacoecon Outcomes Res. 2010;10(5):605–12. Epub 2010/10/19. doi: 10.1586/erp.10.54. PubMed PMID: 20950075.

12. Wilford R, Golden K, Walker DG. Cost-effectiveness of community-based management of acute malnutrition in Malawi. Health Policy Plan. 2012;27(2):127–37. Epub 2011/03/08. doi: 10.1093/heapol/czr017. PubMed PMID: 21378101.

13. Griswold SP, Langlois BK, Shen Y, Cliffer IR, Suri DJ, Walton S, et al. Effectiveness and costeffectiveness of 4 supplementary foods for treating moderate acute malnutrition: results from a cluster-randomized intervention trial in Sierra Leone. The American journal of clinical nutrition. 2021;114(3):973–85. Epub 2021/05/22. doi: 10.1093/ajcn/nqab140. PubMed PMID: 34020452; PubMed Central PMCID: PMCPMC8408853.

14. Kimani-Murage EW, Muthuri SK, Oti SO, Mutua MK, van de Vijver S, Kyobutungi C. Evidence of a Double Burden of Malnutrition in Urban Poor Settings in Nairobi, Kenya. PloS one. 2015;10(6):e0129943. Epub 2015/06/23. doi: 10.1371/journal.pone.0129943. PubMed PMID: 26098561; PubMed Central PMCID: PMCPMC4476587.

15. Concern Worldwide. Nutrition Survey Conducted in the Slums of Nairobi County. Nairobi, Kenya Concern Worldwide 2017.

16. Owino A. Job Groups in Kenya: Civil Servants Salary Nairobi, Kenya2019 [cited 2020 04/09]. Available from: https://www.kenyans.co.ke/news/42378-job-groups-kenya-civil-servants-salary.

17. UNICEF. Ready-to-use therapeutic food (RUTF) price data. Copenhagen: UNICEF, division S; 2022.

18. Salary and Cost of Living Comparison [Internet]. 2022. Available from: http://www.salaryexplorer.com/.

19. Binns P, Myatt M. Does treatment of short or stunted children aged 6-59 months for severe acute malnutrition using ready to use therapeutic food make them overweight? Data from Malawi. Arch Public Health. 2018;76:78. doi: 10.1186/s13690-018-0321-1. PubMed PMID: 30559964; PubMed Central PMCID: PMCPMC6292002.

20. Lelijveld N, Beedle A, Farhikhtah A, Elrayah EE, Bourdaire J, Aburto N. Systematic review of the treatment of moderate acute malnutrition using food products. Maternal & child nutrition. 2020;16(1):e12898. Epub 2019/11/02. doi: 10.1111/mcn.12898. PubMed PMID: 31667981; PubMed Central PMCID: PMCPMC7038867.

21. Nikiema L, Huybregts L, Kolsteren P, Lanou H, Tiendrebeogo S, Bouckaert K, et al. Treating moderate acute malnutrition in first-line health services: an effectiveness cluster-randomized trial in Burkina Faso. The American journal of clinical nutrition. 2014;100(1):241–9. Epub 2014/05/09. doi: 10.3945/ajcn.113.072538. PubMed PMID: 24808482.

22. James P, Sadler K, Wondafrash M, Argaw A, Luo H, Geleta B, et al. Children with Moderate Acute Malnutrition with No Access to Supplementary Feeding Programmes Experience High Rates of Deterioration and No Improvement: Results from a Prospective Cohort Study in Rural Ethiopia. PloS one. 2016;11(4):e0153530. doi: 10.1371/journal.pone.0153530. PubMed PMID: 27100177; PubMed Central PMCID: PMCPMC4839581.

